## Supplementary Materials for "Estimating the replicability of highly cited clinical research (2004-2018)"

| **Highly cited study** | **Effect Measure** | **% of Sample Size** | **Published effect size [95% CI]** | **Effect size with highly cited study [95% CI]** | **Effect size without highly cited study [95% CI]** |
| --- | --- | --- | --- | --- | --- |
| Topalian et al. 2012 (1) | ORR | 8.2 | 0.26 [0.21; 0.31] | 0.24 [0.20; 0.30] | 0.25 [0.19; 0.31] |
| Brahmer et al. 2012 (2) | ORR | 5.5 | 0.27 [0.21; 0.33] | 0.25 [0.19; 0.32] | 0.26 [0.20; 0.33] |
| SYNTAX (3) | OR | 16.7 | 1.42 [1.27; 1.59] | 1.43 [1.27; 1.59] | 1.43 [1.27; 1.61] |
| ACCORD (4) | RR | 17.6 | 0.92 [0.85; 1.00] | 0.92 [0.85; 0.99] | 0.92 [0.85; 0.99] |
| ECASS III (5) | OR | 11.9 | 1.29 [1.16; 1.43] | 1.28 [1.13; 1.45] | 1.37 [1.07; 1.74] |
| MR CLEAN (6) | RR | 17.2 | 1.37 [1.14; 1.64] | 1.47 [1.20; 1.82] | 1.44 [1.15; 1.82] |
| ESCAPE (7) | RR | 10.9 | 1.37 [1.14; 1.64] | 1.47 [1.20; 1.82] | 1.43 [1.14; 1.80] |
| ERSPC (8) | IRR | 24.3 | 0.96 [0.85; 1.08] | 0.98 [0.86; 1.12] | 1.02 [0.93; 1.12] |
| CATIE (9) | HR | 28.6 | 0.68 [0.56; 0.83] | 0.68 [0.55; 0.84] | 0.69 [0.52; 0.91] |
| HERA (10) | HR | 23 | 0.65 [0.55; 0.75] | 0.66 [0.58; 0.76] | 0.62 [0.56; 0.68] |
| EURTAC (11) | HR | 32 | 0.23 [0.17; 0.30] | 0.30 [0.17; 0.53] | 0.26 [0.10; 0.67] |
| EXTEND-IA (12) | OR | 5.60 | 4.04 [2.75; 5.93] | - | - |
| SHARP (13) | HR | 42.1 | 0.69 [0.60; 0.79] | - | - |
| TAXUS-IV (14) | RR | 13.52 | 0.66 [0.59; 0.74] | - | - |
| PROFILE 1007 (15) | HR | 38.68 | 0.46 [0.39; 0.54] | - | - |
| Cheng et al. 2009 (16) | HR | 27.29 | 0.69 [0.54; 0.87] | - | - |

**Table S1. Results from reanalyzed meta-analyses with and without the highly cited study.** Table shows highly cited studies with their effect measures, the % of sample size they account for in the replication meta-analyses, the published results of these meta-analyses, and those of their reanalyses before and after removing the highly cited study. Out of 16 meta-analyses, we were able to reanalyze 11: EXTEND-IA (12) is an individual patient data meta-analysis, whereas the remaining 3 are network meta-analyses with no direct comparisons. One network meta-analysis (17) replicates two highly cited studies ((13) and (16)), making it 4 network meta-analyses that were not possible to conduct independent replications. One network meta-analysis (18) had direct comparisons in its supplementary materials, making it possible to conduct the independent analysis. Differences between the effect sizes of published and reanalyzed meta-analyses using the same studies occur due to changes in meta-analytical methods and software, but are generally small.

**Table S2**

| **Criterion** | **Total** | **Replicated** | **% Replicated [95% CI]** |
| --- | --- | --- | --- |
| Statistical significance | 10 | 8 | 80% [49, 94] |
| 95% CI overlap | 19 | 16 | 84% [62, 94] |
| 95% CI overlap and statistical significance | 19 | 15 | 79% [57, 91] |
| Statistical significance including negative studies | 12 | 8 | 67% [41, 94] |
| Replication estimate within highly cited study’s 95% CI | 19 | 11 | 58% [36, 77] |
| Highly cited study estimate within replication 95% CI | 19 | 9 | 47% [27, 68] |

**Table S2. Replication rates using fully independent replications only.** Rates consider only independent primary studies (i.e. RCTs, phase II trials) and meta-analyses that do not include the highly cited studies. Meta-analyses that could not be reanalyzed were excluded from the analysis. Otherwise, results are displayed in the same way as in **Table 4**.

**Table S3**

| Analysis | All | p-value | Phase 1 trials | RCTs |
| --- | --- | --- | --- | --- |
| Main analysis | 0.99 [0.83 - 1.19] | 0.93 | 1.02 [0.66 - 1.57] | 0.98 [0.79 - 1.20] |
| Publication order | 1.06 [0.89 - 1.27] | 0.48 | 1.02 [0.66 - 1.57] | 1.09 [0.89 - 1.33] |
| Effect coining | 1.04 [0.87 - 1.23] | 0.67 | 1.17 [0.80 - 1.71] | 0.96 [0.79 - 1.18] |
| Publication order + coining | 1.11 [0.94 - 1.31] | 0.21 | 1.17 [0.80 - 1.71] | 1.07 [0.88 - 1.31] |

**Table S3. Effect size inflation using fully independent replications only.** Rates consider only independent primary studies (i.e. RCTs, phase II trials) and meta-analyses that do not include the highly cited studies. Meta-analyses that could not be reanalyzed were excluded from the analysis. Otherwise, results are displayed in the same way as in **Table 5**.

**Table S4.**

| **Predictor** | **Criteria** | **Median (IQR) Replicated** | **Median (IQR) Contradicted** | **p-value** |
| --- | --- | --- | --- | --- |
|  | Significance (p < 0.05) (independent) | 308 [243 - 406] | 441 [338 - 543] | 0.71 |
| Citations/year of the highly cited study | CI overlap [independent] | 337 [247 - 414] | 494 [368 - 570] | 0.36 |
|  | Significance [p < 0.05] & CI overlap [independent] | 346 [269 - 434] | 368 [241 - 532] | 0.81 |
|  | Significance [p < 0.05] [independent] | 2x10^-5^ [4x10^-6^ - 2x10^-3^] | 0.02 [8x10^-3^ - 0.02] | 0.53 |
| p-value of the highly cited study | CI overlap [independent] | 5x10^-4^ [5x10^-5^ - 0.03] | 1x10^-5^ [1x10^-5^ - 1x10^-5^] | 0.83 |
|  | Significance [p < 0.05] & CI overlap [independent] | 3x10^-4^ [5x10^-6^ - 0.03] | 0.02 [8x10^-3^ - 0.02] | 0.91 |
|  | Significance [p < 0.05] [independent] | 659 [408 - 1310] | 182000 [182000 - 182000] | 0.25 |
| Sample size of the highly cited study | CI overlap [independent] | 740 [362 - 2990] | 207 [207 - 207] | 0.36 |
|  | Significance [p < 0.05] & CI overlap [independent] | 659 [316 - 1800] | 91104 [45655 - 136552] | 0.91 |
| **Predictor** | **Criteria** | **# replicated by study design** | **# not replicated by study design** | **p-value** |
| Highly cited study design | CI overlap [independent] | Phase 1 trial: 5/7 RCT: 11/12 | Phase 1 trial: 2/7 RCT: 1/12 | 0.52 |
|  | Significance [p < 0.05] & CI overlap [independent] | Phase 1 trial: 5/7 RCT: 10/12 | Phase 1 trial: 2/7 RCT: 2/12 | 0.60 |
|  | Significance [p < 0.05] [independent] | Pharmacological: 4/5  Other: 4/5 | Pharmacological: 1/5  Other: 1/5 | 1.00 |
| Type of intervention | CI overlap [independent] | Pharmacological: 10/13  Other: 6/6 | Pharmacological: 3/13  Other: 0/6 | 0.52 |
|  | Significance [p < 0.05] & CI overlap [independent] | Pharmacological: 10/13  Other: 5/6 | Pharmacological: 3/13  Other: 1/6 | 1.00 |

**Table S4. Predictors of replicability using fully independent replications only.** Rates consider only independent primary studies (i.e., RCTs, phase II trials) and meta-analyses that do not include the highly cited studies. Meta-analyses that could not be reanalyzed for this purpose were excluded from the analysis. Otherwise, results are displayed in the same way as in **Table 6**. Sample size for each group varies according to the specific criteria: statistical significance: 8 replicated, 2 contradicted; confidence interval overlap: 16 replicated, 3 contradicted; both criteria: 15 replicated, 4 contradicted.

**Figure S1.**

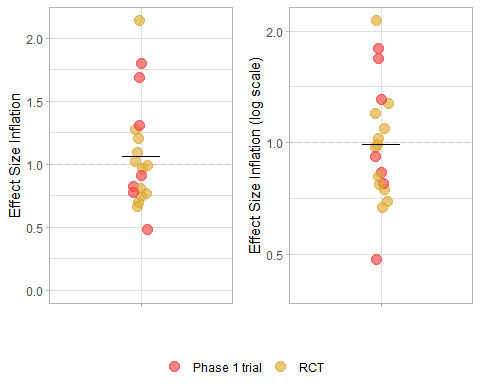

**Figure S1. Inflation of effects using fully independent replications only.** Rates consider only independent primary studies [i.e. RCTs, phase II trials] and meta-analyses that do not include the highly cited studies. Meta-analyses that could not be reanalyzed for this purpose were excluded from the analysis. Otherwise, results are displayed in the same way as in **Figure 3**. Lines indicate the mean of the plotted values (which in the left panel differs from that on **Table S3**, calculated on the basis of log-transformed values).
